# Apparent reductions in COVID-19 Case Fatality Rates reflect changes in average age of those testing positive

**DOI:** 10.1101/2020.09.18.20197160

**Authors:** Alastair Grant

## Abstract

Numbers of COVID-19 infections are rising in many countries, while death and hospitalisation rates remain low. Infection Fatality Rates (IFR) for individual year classes calculated from seroprevalence data and the ages of those dying in England show a very strong log-linear relationship with age, and allow us to predict fatality rates for those testing positive on each day based on their ages. Since the peak of the epidemic, reductions in the ages of cases account for an eight fold fall in fatality rates. Over the same period, increased testing intensity appears to have increased infections detected amongst the most vulnerable by a factor of at least five, and between 15 and 24 fold in the population as a whole. Together these two factors are sufficient to explain the large observed change in the ratio of deaths to reported cases. We can also use these methods to give a more precise early warning system of future increases in mortality rates than raw case numbers. Although case numbers are currently increasing markedly, a continuing reduction in numbers of older individuals being infected means that the predicted increase in mortality rates is much slower.

## Introduction

Numbers of reported cases of COVID-19 have been increasing in the UK and several other European countries, while death rates have been stable (ECDC, 2020). This has resulted in an apparent reduction in case fatality rates (CFR). Possible reasons include improved clinical outcomes for severe cases; reduced virulence of the pathogen; a reduction of the proportion of cases amongst the elderly and those with pre-existing conditions where mortality rates are higher and increased testing leading to the inclusion of individuals with mild or no symptoms in the case count and (Vaughan, 2020).

Calculating the true infection fatality rate (IFR, as opposed to CFR) requires estimation of numbers who were infected or died without having a positive test. We sought evidence on changing demographics in those infected and how this might explain apparent differences in CFR and IFR. We used data on COVID-19 seroprevalence in England and the counts and ages of deaths attributed to COVID-19 both with and without a positive COVID-19 test to estimate age-specific IFR. The resulting rates, including values for individual year groups between 20 and 90, can be combined with data on the ages of those testing positive to assess the proportion of the reduction in CFR likely to be due to the substantial decline in the average age of confirmed cases since the peak of the epidemic in April 2020 and to predict whether recent rises in case numbers will lead to similar mortality increases.

## Community seroprevalence surveys

Systematic seroprevalence testing suggests “a trend towards decreasing percentage of individuals testing positive for antibodies with increasing age” (ONS, 2020a; Ward et al., 2020). Multiplying age-specific seroprevalence by numbers in each age class (ONS, 2020b) gives estimates for the cumulative numbers of individuals in each age group who have had COVID-19. Overall totals for England are 3.1 and 3.6 million for the ONS and React2 seroprevalence data. This compares with a cumulative total of 302 175 confirmed cases up until 7 Sept 2020 (UK Government, 2020). The actual count of individuals who have been infected will be higher, as approximately 9% of those known to have been infected are not seropositive to standard tests (Gudbjartsson et al., 2020).

## Case numbers and infection fatality rates

Anonymised linelists of individuals testing positive and deaths attributed to COVID-19 in England, including individual ages, were obtained from Public Health England (PHE) on 7 Sept 2020, reflecting data reported up until 6 September 2020. Estimates of the number who have died from COVID-19 through 6 September 2020 range from 36 896 (UK Government, 2020, counting only those who have died within 28 days of a positive test) to 56 950 (all deaths in PHE daily data file, including all mortality subsequent to a positive test and deaths attributed to COVID-19 but without a positive test). Combining the low and high estimates of numbers of deaths and of total infections gives overall values for IFR ranging from 1.03% to 1.84%. It is acknowledged (Raleigh, 2020) that some of the excess deaths which occurred early in the pandemic, especially of the elderly in residential care homes, were also due to COVID-19, but no attempt was made to include these in our analysis.

## Age-specific infection fatality rates

We can calculate age-specific IFRs by dividing the number of deaths in each year class by the estimated number of cases. IFR shows a log-linear relationship with age (Fig. 1b, c.f. Levin, Meyerowitz-Katz, Owusu-Boaitey, Cochran, & Walsh, 2020; Spiegelhalter, 2020), with IFR approximately doubling every 6 years. There are small differences in the shape of the relationship and larger differences in the absolute values of IFR (varying from 25-42% for individuals aged 90+) depending upon the seropositivity and death data used but after excluding individuals under 20, R^2^ is greater than 0.99 for all four combinations.

## Numbers and ages of individuals testing positive

The number of individuals testing positive for COVID-19 in England peaked at a seven day average of 4140 in week ending 24 April 20, fell to 535 in week ending 4 July and rose to 1584 in week ending 5^th^ September (Fig 2a). The average age of people testing positive rose during the early part of the epidemic, but has since steadily decreased (Fig. 2b). The age-specific relationship shown in Figure 1 gives us an IFR for each individual, from which we can calculate an expected overall IFR and number of deaths, given the age distribution of those testing positive. Predicted fatality rate of those testing positive reached a peak of 6.0% in April, falling to 0.78% for the most recent week (Fig. 2c), an 8 fold reduction. Predicted number of deaths reached a minimum of 8 for those testing positive in week ending 22^nd^ August, rising to 11 a day for tests carried out in week ending 6^th^ September (compared to an observed average of 8 or fewer daily deaths throughout the period from week ending 30 Aug 2020 until 6^th^ September 2020). Predicted deaths are only 176 at the peak (Fig 2d), compared with a maximum daily rate in England of 974 on 8^th^ April and a 7 day rolling average of 876 for the week ending 12^th^ April (Fig. 1a; UK Government, 2020). However, only 9 390 tests per day were administered in week ending 4 April and 14 560 a day in week ending 24 April, compared with 134 997 in week ending 30^th^ Aug 2020 (UK Government, 2020). The underestimation of mortality rates at the peak presumably reflects the relatively low testing intensity then in operation. It also suggests that counting only individuals who have died following a positive test is likely to substantially underestimate numbers of deaths resulting from COVID during the peak of the epidemic.

**Figure 1.**
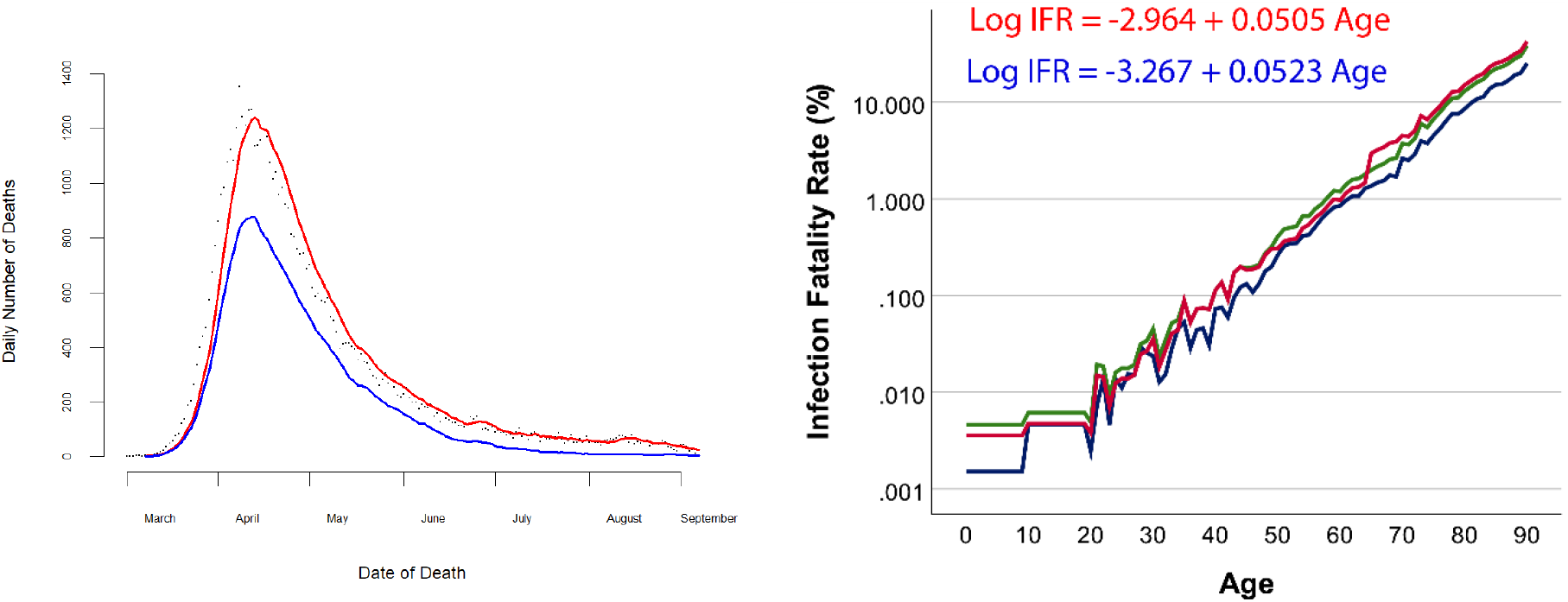
Deaths in England attributed to COVID-19. a) all deaths in Public Health England linelist file attributed to COVID-19 (dots) plus 7 day rolling mean (red line) and 7 day rolling mean of numbers dying within 28 days of a positive test (blue line). b) Estimated Infection Fatality Rate against age. Values for ages 0-9 and 10-19 years are averaged into one category as the numbers are small. Individuals age 90+ are similarly grouped together. Green and blue indicate ONS seropositivity survey combined with all deaths and deaths within 28 days of a positive test respectively. Red indicates React2 seropositivity survey combined with all deaths.

**Figure 2.**
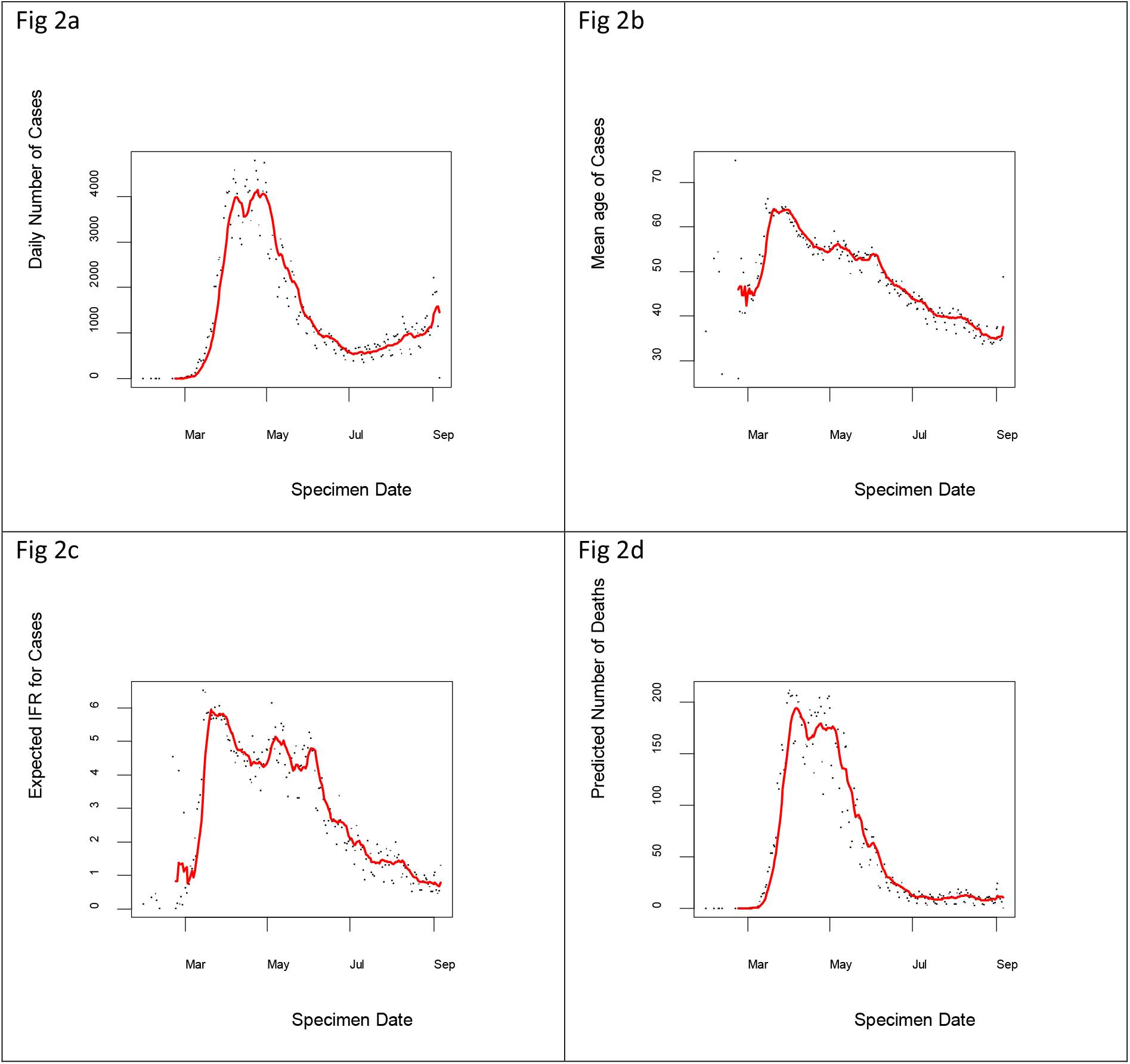
a) number of positive tests against time, and 7 day rolling mean (red line); b) Average age of individuals testing positive; c) Expected infection fatality rate for individuals testing positive each day, given their distribution of ages; d) Number of deaths per day predicted from ages and numbers of those testing positive.

## Discussion

At the epidemic peak there were just over 4000 positive tests each day in England and an average daily death rate of nearly 900. At the end of the period studied here, there were fewer than 10 deaths but over 1500 positive tests each day. Our analysis shows that reducing age of cases makes a substantial contribution to this change, accounting for an approximate 8 fold reduction in predicted IFR *for those who are testing positive*. The large increase in testing intensity over the same period will also contribute, as an individual who is infected is much more likely to be tested now than at the peak of the epidemic. For the April peak period, the difference between actual and predicted daily death rates suggests that testing at the current intensity would have detected at least five times as many positive cases (around 20 000 per day). This assumes that the age distribution of infected individuals then matched that of those who tested positive. However, testing was focussed in hospitals in early April so was likely to be biased towards those with severe symptoms who are more likely to be elderly. If we estimate numbers of infections across the population as a whole by combining numbers of deaths with the two IFR estimates for the whole period of the pandemic, then the number of infections may have peaked at between 53 and 95 thousand per day – between 15 and 24 times higher than the number of cases detected at the time.

## Data Availability

The anonymised linelist data is available for research purposes from Public Health England (https://www.gov.uk/government/organisations/public-health-england), subject to completion of an Non Disclosure Agreement

## Acknowledgements

We are grateful to Public Health England for providing access to the anonymised case and mortality line list files and to Paul Hunter and Julii Brainard for comments on the manuscript.

